# Characterisation of the neonatal brain using myelin-sensitive magnetisation transfer imaging

**DOI:** 10.1101/2023.02.01.23285326

**Authors:** Manuel Blesa Cábeza, Kadi Vaher, Elizabeth N. York, Paola Galdi, Gemma Sullivan, David Q. Stoye, Jill Hall, Amy E. Corrigan, Alan J. Quigley, Adam D. Waldman, Mark E. Bastin, Michael J. Thrippleton, James P. Boardman

## Abstract

A cardinal feature of the encephalopathy of prematurity is dysmaturation of developing white matter and subsequent hypomyelination. Magnetisation transfer imaging (MTI) offers surrogate markers for myelination including magnetisation transfer ratio (MTR) and magnetisation transfer saturation (MTsat). Using data from 105 neonates, we characterise MTR and MTsat in the developing brain and investigate how these markers are affected by gestational age at scan and preterm birth. We explore correlations of the two measures with fractional anisotropy (FA), radial diffusivity (RD) and T1w/T2w ratio which are commonly used markers of white matter integrity in early life. We used two complementary analysis methods: voxel-wise analysis across the white matter skeleton, and tract-of-interest analysis across 16 major white matter tracts. We found that MTR and MTsat positively correlate with gestational age at scan. Preterm infants at term-equivalent age had lower values of MTsat in the genu and splenium of the corpus callosum, while MTR was higher in central white matter regions, the corticospinal tract and the uncinate fasciculus. Correlations of MTI metrics with other MRI parameters revealed that there were moderate positive correlations between T1w/T2w and MTsat and MTR at voxel-level, but at tract-level FA had stronger positive correlations with these metrics. RD had the strongest correlations with MTI metrics, particularly with MTsat in major white matter tracts. The observed changes in MTI metrics are consistent with an increase in myelin density during early postnatal life, and lower myelination and cellular/axonal density in preterm infants at term-equivalent age compared to term controls. Furthermore, correlations between MTI-derived features and conventional measures from dMRI provide new understanding about the contribution of myelination to non-specific imaging metrics that are often used to characterise early brain development.

## 1 Introduction

The integrity of brain development during pregnancy and the new born period is critical for life-long cognitive function and brain health. During the second and third trimesters of pregnancy, there is a phase of rapid brain maturation characterised by volumetric growth, increases in cortical complexity, white matter (WM) organisation and myelination (Counsell et al., 2019; Dubois et al., 2021). Early exposure to extrauterine life due to preterm birth, defined as birth *<* 37 weeks of gestation, affects around 11% of births and is closely associated with neurodevelopmental, cognitive and psychiatric impairment (Johnson and Marlow, 2017; Nosarti et al., 2012; Wolke et al., 2019), and alterations to brain development that are apparent using MRI (Boardman and Counsell, 2019; Counsell et al., 2019; Pecheva et al., 2018).

Structural MRI (T1- and T2-weighted) and diffusion MRI (dMRI) have revealed a phenotype of preterm birth that includes changes in global and regional tissue volumes and cortical complexity, and altered microstructural integrity of the WM (Counsell et al., 2019; Pecheva et al., 2018). These imaging features capture the encephalopathy of prematurity (EoP), which is thought to underlie long term impairments (Volpe, 2009). Diffusion metrics are influenced by microstructural properties of the underlying tissue including axonal density and diameter, and water content; although myelination may alter/contribute to water diffusivity, myelin does not directly contribute to the diffusion signal due to short T2 (Mancini et al., 2020; van der Weijden et al., 2020).

Pre-oligodendrocytes are particularly vulnerable to hypoxia-ischaemia and inflammation associated with preterm birth (Back and Volpe, 2018; Volpe et al., 2011). Although this cell population is mostly replenished following primary injury, subsequent differentiation into myelin-producing oligodendrocytes can fail, leading to hypomyelination (Billiards et al., 2008; Volpe, 2019). Therefore, imaging tools that more specifically model myelination in early life could enhance biology-informed assessment of EoP.

Several MRI techniques are sensitive to myelin content (Lazari and Lipp, 2021; Mancini et al., 2020; Piredda et al., 2021). In the developing brain, the most commonly applied myelin-sensitive imaging techniques are those based on relaxometry, such as T1 (or its inverse, R1) or T2 (or its inverse, R2) mapping (e.g. Counsell et al., 2003; Grotheer et al., 2022; Kulikova et al., 2015; Leppert et al., 2009; Maitre et al., 2014; Schneider et al., 2016), quantification of myelin water fraction (e.g. Dean et al., 2014; Deoni et al., 2011; Melbourne et al., 2013), and calculation of T1w/T2w ratio (e.g. Filimonova et al., 2023; Grotheer et al., 2023; Lee et al., 2015; Soun et al., 2017). However, T1 and T2 relaxation are partly determined by iron concentration (Birkl et al., 2019; Stüber et al., 2014), and T1w/T2w ratio correlations with other myelin-sensitive MRI parameters and histological myelin measurements are low (Arshad et al., 2017; Sandrone et al., 2023; Uddin et al., 2018).

Magnetisation transfer imaging (MTI) is a family of MRI techniques sensitive to subtle pathological changes in tissue microstructure which cannot typically be quantified with conventional MRI (Sled, 2018). MTI is based on the exchange of magnetisation between immobile protons associated with macromolecules, and mobile protons in free water. MTI is sensitive to myelin-associated macromolecules such as cholesterol, myelin basic protein, sphingomyelin and galactocerebrosides, and thus it provides a surrogate marker of myelin integrity (Mancini et al., 2020). To date, MTI has mainly been used to study demyelinating diseases such as multiple sclerosis (Sled, 2018; York et al., 2022b).

Magnetisation transfer ratio (MTR), calculated as the percentage change in signal with and without off-resonance radiofrequency saturation, is the most widely used MTI metric. MTR is, however, susceptible to transmit (B1^+^) field inhomogeneities (Helms et al., 2010a) and T1 relaxation effects, and varies widely depending upon specific acquisition parameters (Samson et al., 2006; York et al., 2022c). Biological interpretation of MTR is therefore challenging, which presents a barrier to clinical translation. The addition of a T1w sequence allows computation of magnetisation transfer saturation (MTsat) which inherently corrects for B1^+^ inhomogeneities and T1 relaxation to a substantial degree (Helms et al., 2008b; Samson et al., 2006). MTsat hence addresses some limitations of MTR within clinically feasible acquisition times and the resulting parametric maps have visibly better tissue contrast compared with MTR (Helms et al., 2008b; Samson et al., 2006; York et al., 2022b). Higher values of MTR and MTsat are associated with greater myelin density.

In neonates, MTR has been used to characterise brain development during the preterm period from birth up to term-equivalent age (TEA): in general, MTR values in WM increase with gestational age (GA) at scan (Nossin-Manor et al., 2015, 2013, 2012; Zheng et al., 2016). In addition, at the age of 4 years, children born very preterm have lower MTR values across the WM compared to term-born peers, and WM MTR positively correlates with language and visuo-motor skills (Vandewouw et al., 2019). Furthermore, in infancy, an MTI-derived macromolecular proton fraction (MPF) has predictive value for neurocognitive outcomes (Corrigan et al., 2022; Zhao et al., 2022). Yet, the use of MTI in the neonatal brain has been scarce, and, to the best of our knowledge, MTsat has not been used to study myelination in human neonates. Furthermore, no studies have explored the effect of preterm birth on MTI metrics in comparison to term controls at TEA.

In this work, we aimed to obtain a description of brain myelination processes by applying MTI in the neonatal period. We had three objectives: 1) to characterise the associations of MTsat and MTR in neonatal WM with GA at MRI scan; 2) to test the hypothesis that myelin-sensitive features would differ between preterm infants at TEA and term controls; and 3) to assess the relationship between MTI metrics and the T1w/T2w ratio, a commonly used myelin proxy, fractional anisotropy (FA), which is most robustly associated with EoP but is not specific to myelination, and radial diffusivity (RD), a diffusion biomarker that has been related to myelin pathologies (Lazari and Lipp, 2021; Mancini et al., 2020; Song et al., 2002).

## 2 Material and methods

### 2.1 Participants and data acquisition

Participants were very preterm infants (GA at birth < 32 completed weeks) and term-born controls recruited as part of a longitudinal study designed to investigate the effects of preterm birth on brain structure and long-term outcome (Boardman et al., 2020). The cohort exclusion criteria were major congenital malformations, chromosomal abnormalities, congenital infection, overt parenchymal lesions (cystic periventricular leukomalacia, haemorrhagic parenchymal infarction) or post-haemorrhagic ventricular dilatation. The study was conducted according to the principles of the Declaration of Helsinki, and ethical approval was obtained from the UK National Research Ethics Service. Parents provided written informed consent. 105 neonates (83 preterm and 22 term) who underwent MTI at TEA at the Edinburgh Imaging Facility (Royal Infirmary of Edinburgh, University of Edinburgh, UK) were included in the current study.

A Siemens MAGNETOM Prisma 3 T clinical MRI system (Siemens Healthcare Erlangen, Germany) and 16-channel phased-array paediatric head coil were used to acquire a three-dimensional (3D) T1w magnetisation prepared rapid gradient echo (MPRAGE) structural image (voxel size = 1 mm isotropic, echo time [TE] = 4.69 ms and repetition time [TR] = 1970 ms); 3D T2-weighted SPACE images (T2w) (voxel size = 1mm isotropic, TE = 409 ms and TR = 3200 ms) and axial dMRI data. dMRI volumes were acquired in two separate acquisitions to reduce the time needed to re-acquire any data lost to motion artifacts: the first acquisition consisted of 8 baseline volumes (b = 0 s/mm^2^ [b0]) and 64 volumes with b = 750 s/mm^2^; the second consisted of 8 b0, 3 volumes with b = 200 s/mm^2^, 6 volumes with b = 500 s/mm^2^ and 64 volumes with b = 2500 s/mm^2^. An optimal angular coverage for the sampling scheme was applied (Caruyer et al., 2013). In addition, an acquisition of 3 b0 volumes with an inverse phase encoding direction was performed. All dMRI volumes were acquired using single-shot spin-echo echo planar imaging (EPI) with 2-fold simultaneous multi-slice and 2-fold in-plane parallel imaging acceleration and 2 mm isotropic voxels; all three diffusion acquisitions had the same parameters (TR/TE 3400/78.0 ms). Acquisitions affected by motion artifacts were re-acquired multiple times as required; dMRI acquisitions were repeated if signal loss was seen in 3 or more volumes. MTI consisted of three sagittal 3D multi-echo spoiled gradient echo images (TE = 1.54/4.55/8.56 ms, 2 mm isotropic acquired resolution): magnetisation transfer (TR = 75 ms, flip angle = 5°, gaussian MT pulse (offset 1200 Hz, duration 9.984 ms, flip angle = 500°) [MT_on_]), proton density-weighted (TR = 75 ms, flip angle = 5° [MT_off_]) and T1w (TR = 15 ms, flip angle = 14° [MT_T1w_]) acquisitions. All acquisition parameters are detailed in the cohort protocol (Boardman et al., 2020).

Infants were fed and wrapped and allowed to sleep naturally in the scanner. Pulse oximetry, electrocardiography and temperature were monitored. Flexible earplugs and neonatal earmuffs (MiniMuffs, Natus) were used for acoustic protection. All scans were supervised by a doctor or nurse trained in neonatal resuscitation.

### 2.2 Data preprocessing

The image analysis was performed with MRtrix3 (Tournier et al., 2019), FSL 5.0.11 (Smith et al., 2004), ANTs (Avants et al., 2008), the developing Human Connectome Project (dHCP) pipeline (Makropoulos et al., 2018) and MATLAB R2022a.

dMRI processing was performed as follows: for each subject, the two dMRI acquisitions were first concatenated and then denoised using a Marchenko-Pastur-PCA-based algorithm (Veraart et al., 2016); eddy current, head movement and EPI geometric distortions were corrected using outlier replacement and slice-to-volume registration (Andersson et al., 2017, 2016, 2003; Andersson and Sotiropoulos, 2016); bias field inhomogeneity correction was performed by calculating the bias field of the mean b0 volume and applying the correction to all the volumes (Tustison et al., 2010). The DTI model was fitted in each voxel using the weighted least-squares method *dtifit* as implemented in FSL using only the b = 750 s/mm^2^ shell.

Structural MRI (T1w and T2w) images were processed using the dHCP minimal processing pipeline to obtain the bias field corrected and coregistered T2w and T1w, brain masks, tissue segmentation and the different tissue probability maps (Makropoulos et al., 2018, 2014). Then T1w/T2w ratio maps were obtained using the bias field corrected images. The T1w/T2w maps were edited to remove voxels with intensities higher than the mean + 5 standard deviations. Note that a calibration step was not included, as the full dataset was scanned with the same parameters in the same scanner, minimising differences in intensity scale (Ganzetti et al., 2014).

### 2.3 Magnetisation transfer imaging processing

MTI data were processed as previously described (York et al., 2022b, 2020). The three echoes were summed together to increase the signal-to-noise ratio (SNR) (Helms and Hagberg, 2009) for each MT image (MT_off_, MT_on_ and MT_T1w_). MT_on_ and MT_T1w_ images were coregistered to the MT_off_ image using flirt. From (Helms et al., 2010b, 2008a) we can define the amplitude of the spoiled gradient echo at the echo time (App) as:

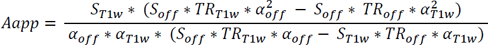

where *S*, *TR* and *α* represent the signal intensity, the repetition time (in seconds), and the flip angle (in radians), respectively. The subscript *off* stands for the proton density-weighted acquisition and the subscript *T1w* for T1-weighted image.

The R1app is expressed as:

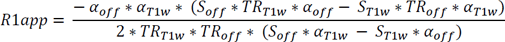

By combining R1app and Aapp, we can obtain the MTsat:

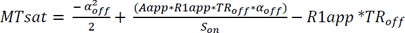

where *S_on_* represents the intensity signal of the magnetization transfer weighted image. Finally, the MTR can be obtained as follows:

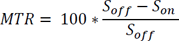

### 2.4 Registration to a common space

MTsat and MTR maps were registered to the structural T1w (MPRAGE) images processed with the dHCP pipeline using ANTs symmetric normalisation (SyN) (Avants et al., 2008). The tissue probability maps for the grey matter (GM) and WM were obtained from the dHCP pipeline (Makropoulos et al., 2018). Nonlinear diffeomorphic multimodal registration was then performed between age-matched T2w and GM/WM tissue probability maps from the dHCP extended volumetric atlas (Fitzgibbon et al., 2020; Schuh et al., 2018) to the subjects T2w and GM/WM tissue probability using SyN (Avants et al., 2008). This was combined with the corresponding template-to-template transformation to yield a structural-to-template (40 weeks GA) transformation, which was finally combined with the MTsat-to-structural transformation to obtain the final MTsat-to-template alignment. By combining all the transformations, image registration was performed with only one interpolation step.

### 2.5 Tract-based spatial statistics

The mean b0 EPI volume of each subject was co-registered to their structural T2w volume using boundary-based registration (Greve and Fischl, 2009). This was combined with the structural-to-template transformation to create the diffusion-to-template transformation and propagate the FA maps to the template space.

The FA maps were averaged and used to create the skeleton mask. MTsat and MTR parametric maps were propagated to template space using the MTsat-to-template transformation and projected onto the skeleton (Smith et al., 2006).

### 2.6 White matter tracts of interest

Sixteen WM tracts were generated in each subject’s diffusion space as previously described (Vaher et al., 2022). Briefly, the tract masks were propagated from the ENA50 atlas (Blesa et al., 2020). These masks were used as a set of regions of interest (ROI) for seeding the tractography, creating the tracts in native diffusion space. Then, the tracts were binarised only including voxels containing at least 10% of the tracts and propagated to MTsat space by combining the MTsat-to-structural and diffusion-to-structural transformations to calculate the mean values in each tract.

### 2.7 Statistical analysis

Tract-based statistical analyses were conducted in R (version 4.0.5) (R Core Team, 2022). We performed multivariate multiple linear regression analyses for all WM tracts, with the tract-average metric as the outcome and preterm status and GA at scan as the predictor variables. The outcome variables as well as GA at scan were scaled (z-transformed) before fitting the models, thus, the regression coefficients reported are in the units of standard deviations. P-values were adjusted for the false discovery rate (FDR) using the Benjamini-Hochberg procedure (Benjamini and Hochberg, 1995) across all MTI metrics separately for the effects of preterm birth and GA at scan; and independently for the comparative FA, RD and T1w/T2w ratio tract-based analyses. The WM tract results were visualised using ParaView (ParaView Developers, 2020), with standardised betas represented as the effect size.

Voxel-wise statistical analysis was performed using a general linear univariate model with PALM (Winkler et al., 2014). Two different contrasts were tested: correlation with GA at scan adjusting for preterm status, and term vs preterm comparison adjusting for GA at scan. Family-wise error correction (FWER), across modalities for MTI metrics (MTsat and MTR) and separately for the complementary FA, RD and T1w/T2w analyses (Winkler et al., 2016), and threshold-free cluster enhancement (TFCE) were applied with a significance level of p*<*0.05 (Smith and Nichols, 2009).

The distributions of MTI metrics, FA, RD and T1w/T2w ratio were compared using two-dimensional histograms of co-registered indexed voxels (RNifti and ggplot2::geom bin2d packages in R) (York et al., 2022a) and voxel-wise correlation analyses between the metrics were performed with repeated measures correlation as implemented in the R package *rmcorr* (Bakdash and Marusich, 2017); this was performed in the WM tissue segmentation obtained from the dHCP pipeline (Makropoulos et al., 2018). Tract-wise correlation coefficients were calculated using Pearson’s r. The average Pearson’s correlation coefficient across all tracts was calculated by first transforming the Pearson’s r values to Fisher’s Z, taking the average, and then back-transforming the value to Pearson’s correlation coefficient (Corey et al., 1998).

### 2.8 Data and code availability

Requests for anonymised data will be considered under the study’s Data Access and Collaboration policy and governance process (https://www.ed.ac.uk/centre-reproductive-health/tebc/about-tebc/for-researchers/data-access-collaboration). The scripts for the data analysis in this paper are available here: https://git.ecdf.ed.ac.uk/jbrl/neonatal-mtsat.

## 3 Results

### 3.1 Sample characteristics

The study group consisted of 105 neonates: 83 participants were preterm and 22 were term-born controls. Participant characteristics are provided in Table 1. Among the preterm infants, 15 (18.1%) had bronchopulmonary dysplasia (defined as need for supplementary oxygen ≥ 36 weeks GA), 3 (3.6%) developed necrotising enterocolitis requiring medical or surgical treatment, 15 (18.1%) had one or more episodes of postnatal sepsis (defined as detection of a bacterial pathogen from blood culture, or physician decision to treat with antibiotics for ≥ 5 days in the context of growth of coagulase negative *Staphylococcus* from blood or a negative culture but raised inflammatory markers in blood), and 2 (2.4%) required treatment for retinopathy of prematurity.

**Table 1:**
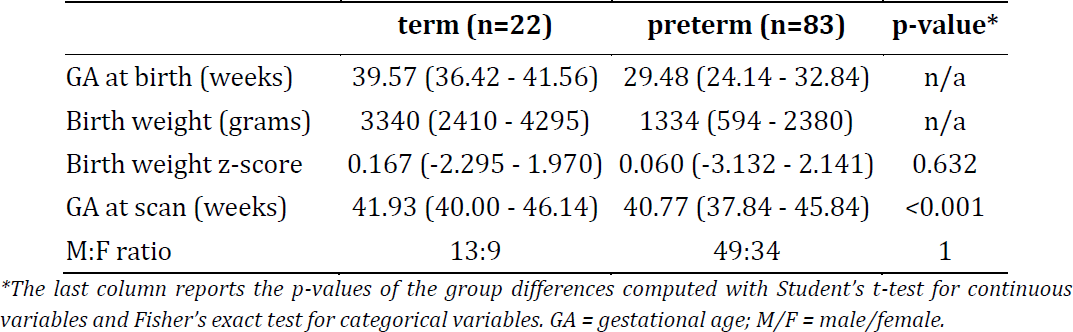
Neonatal participant characteristics.

|  | term (n=22) | preterm (n=83) | p-value* |
| --- | --- | --- | --- |
| GA at birth (weeks) | 39.57 (36.42 - 41.56) | 29.48 (24.14 - 32.84) | n/a |
| Birth weight (grams) | 3340 (2410 - 4295) | 1334 (594 - 2380) | n/a |
| Birth weight z-score | 0.167 (-2.295 - 1.970) | 0.060 (-3.132 - 2.141) | 0.632 |
| GA at scan (weeks) | 41.93 (40.00 - 46.14) | 40.77 (37.84 - 45.84) | <0.001 |
| M:F ratio | 13:9 | 49:34 | 1 |

\*The last column reports the p-values of the group differences computed with Student's t-test for continuous variables and Fisher's exact test for categorical variables. GA = gestational age; M/F = male/female.

### 3.2 Magnetisation transfer imaging metrics in association with gestational age at scan and preterm birth

The average MTsat and MTR maps for the term and preterm infants are shown in Figure 1A. From visual inspection of the averaged maps, MTsat and MTR show similar values across the two groups, although preterm infants at TEA have lower MTsat values mostly in the frontal regions and higher MTR values in the central regions. Tract-averaged values for MTsat and MTR for term and preterm groups are provided in Supplementary Table 1 and visualised in Figure 1B. The highest MTsat values are observed in the corticospinal tract, whilst MTR is highest in the anterior thalamic radiation and cingulum cingulate, followed by the corticospinal tract. The lowest values for MTsat and MTR are observed in the inferior longitudinal fasciculus.

**Figure 1:**
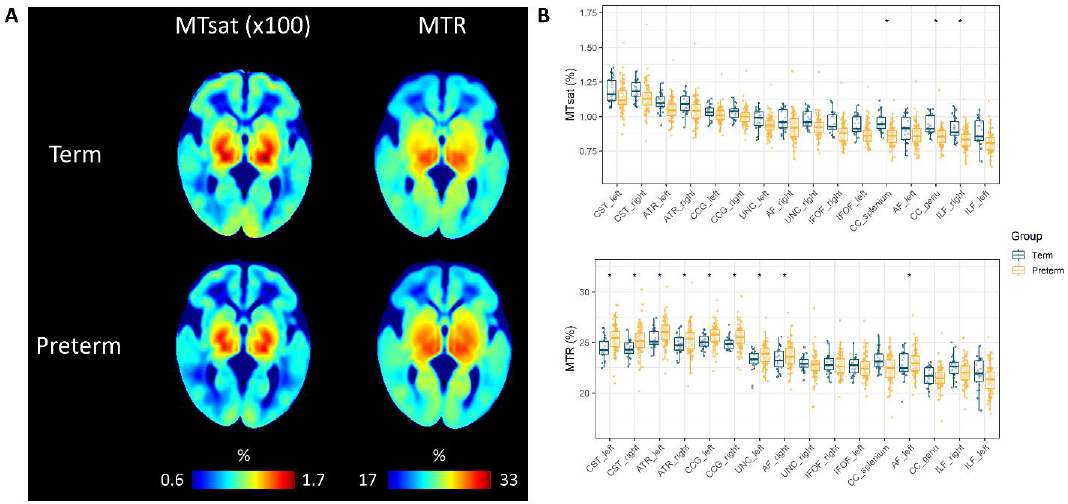
(A) Neonatal MTsat and MTR maps averaged across term and preterm subjects in the study. (B) Tract-averaged MTI metrics in the 16 white matter tracts; tracts are ordered by the values of MTsat. Asterisks (*) indicate tracts with statistically significantly different values between term and preterm infants. MTR = magnetisation transfer ratio, MTsat = magnetisation transfer saturation, CC genu = corpus callosum genu/forceps minor, CC splenium = corpus callosum splenium/forceps major, CST = corticospinal tract, IFOF = inferior fronto-occipital fasciculus, ILF = inferior longitudinal fasciculus, AF = arcuate fasciculus, UNC = uncinate fasciculus, CCG = cingulum cingulate gyrus, ATR = anterior thalamic radiation.

We used two complementary approaches to study the effect of GA at scan and preterm birth on the MTI metrics: voxel-wise in the WM skeleton, and ROI-based using mean values in 16 major WM tracts (Vaher et al., 2022).

MTsat and MTR are positively correlated with GA at scan within the neonatal period between 37-46 weeks of gestation after adjusting for preterm birth. These results were visible on both voxel-wise (Figure 2 left panel) and tract-based analyses (Figure 3 left panel). Positive correlations for both MTsat and MTR with GA at scan were observed when assessed separately in term and preterm groups (Supplementary Figure 1; Supplementary Tables 2-3).

**Figure 2:**
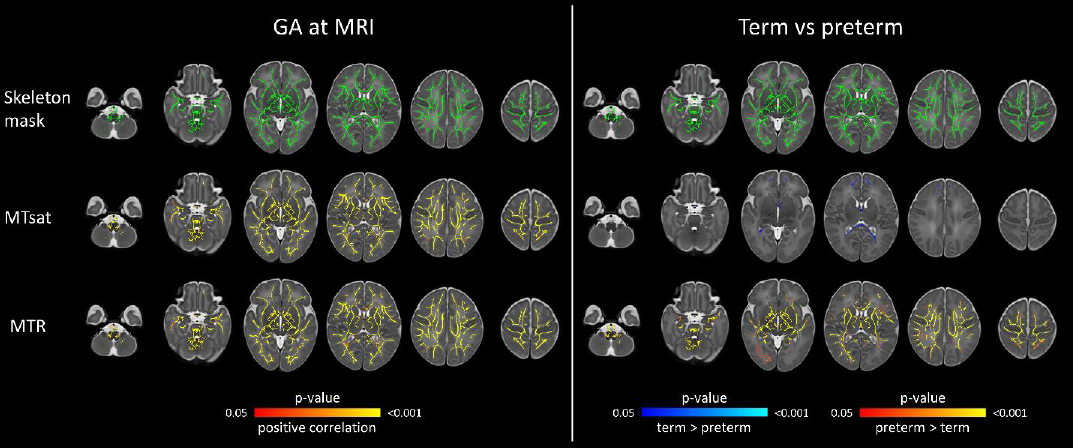
Voxel-wise analysis showing effects of GA at scan and preterm birth on magnetisation transfer imaging metrics. Models were mutually adjusted for GA at scan and preterm status. The first row represents the WM skeleton mask (green) where voxel values were compared. In left panel, voxels that have positive correlation with GA at scan are indicated in red-yellow. In right panel, voxels that have higher values in preterm compared with term group are indicated in red-yellow; voxels that have higher values in term compared with preterm group are indicated in blue-light blue. Overlaid on the dHCP T2w 40-week template. Results are reported after 5000 permutations, p-values corrected using TFCE and FWE with a significance level of p<0.05. For visualisation: anatomic left is on the right side of the image. GA = gestational age, MTR = magnetisation transfer ratio, MTsat = magnetisation transfer saturation, FWE = family-wise error correction, TFCE = threshold-free cluster enhancement.

**Figure 3:**
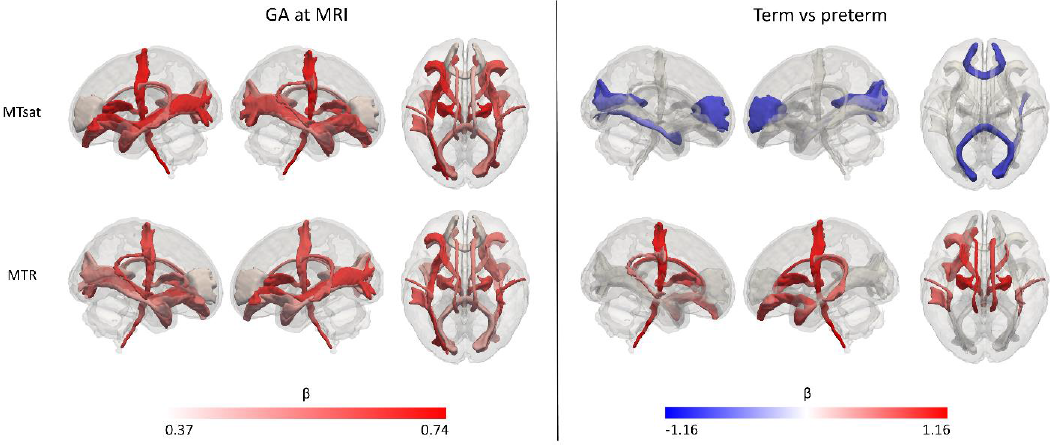
Results of the white matter tract-based analysis of magnetisation transfer imaging metrics at term-equivalent age showing the effects of GA at MRI (left panel) and differences in preterm infants versus term-born controls (right panel). Effect sizes are represented as standardised beta coefficients from the multiple regression where white matter tract values were the outcomes and preterm status and GA at scan the predictors; only statistically significant tracts (FDR-corrected across the two modalities) are coloured. Colour map for the effect sizes was calculated separately for the effects of preterm birth and GA at scan. In left panel, red indicates positive correlation with GA at MRI. In right panel, blue indicates higher values in term infants and red indicates higher values in preterm infants. GA = gestational age, MTR = magnetisation transfer ratio, MTsat = magnetisation transfer saturation.

Complementary analyses for DTI metrics showed positive correlations for FA and negative for RD with GA at scan across the WM skeleton (Supplementary Figure 2 left panel) and tracts (Supplementary Table 4). T1w/T2w ratio had statistically significant positive correlations with GA at scan in the majority of tracts, except in the arcuate fasciculus, corpus callosum and cingulum cingulate (Supplementary Table 4; Supplementary Figure 2 left panel). On average, GA at scan correlations with MTsat, MTR, FA and RD were of a similar magnitude (mean β range across tracts |0.524| - |0.589|), whilst correlation with T1w/T2w ratio was lower (mean β = 0.197).

Although we observed that both MTsat and MTR positively correlate with GA at scan, the effect of preterm birth was different for these two metrics. Compared to preterm infants, term infants had higher MTsat values in the genu and splenium of the corpus callosum (Figure 2 right panel). Tract-level analyses showed similar results (Figure 3 right panel). In contrast, MTR was higher in preterm infants, with significant differences in the central WM regions, and in the corticospinal tracts and uncinate fasciculi (Figure 2 and 3 right panels).

Complementary analysis of DTI metrics (Supplementary Figure 2 right panel, Supplementary Table 4) showed higher FA values in the term group, with the strongest effects observed in the genu and splenium of the corpus callosum and the uncinate. These higher values of FA in the term group were paralleled with lower values of RD. This accords with our previous findings in the wider cohort (Vaher et al., 2022). T1w/T2w ratio was significantly higher in the term group across the WM skeleton and the tracts (Supplementary Figure 2 right panel, Supplementary Table 4), with a large effect size (mean β = |0.695|).

### 3.3 Correlation between MTI metrics, T1w/T2w ratio, FA and RD

We performed both voxel-wise (Figure 4) and tract-wise correlations (Figure 5) of T1w/T2w ratio, FA and RD with the MTI-derived metrics. At the voxel-wise level (Figure 4), T1w/T2w ratio had moderate positive correlations with MTsat (r = 0.446) and MTR (r = 0.328); on the other hand, FA only has weak positive correlation with MTR (r = 0.258) and very weak correlations with MTsat (r = 0.161). However, these correlation trends are different at the tract level (Figure 5; Supplementary Table 5) where FA shows strong positive correlation with MTsat (mean r = 0.646) and moderate correlation with MTR (mean r = 0.512). However, T1w/T2w ratio shows much weaker correlations (mean r = 0.257 with MTsat and mean r = 0.254 with MTR). RD had relatively stronger negative correlations with MTsat and MTR at the voxel-wise level (r = −0.595 and r = −0.597, respectively) and even stronger in individual white matter tracts (mean r = −0.799 and mean r = −0.605, respectively).

**Figure 4:**
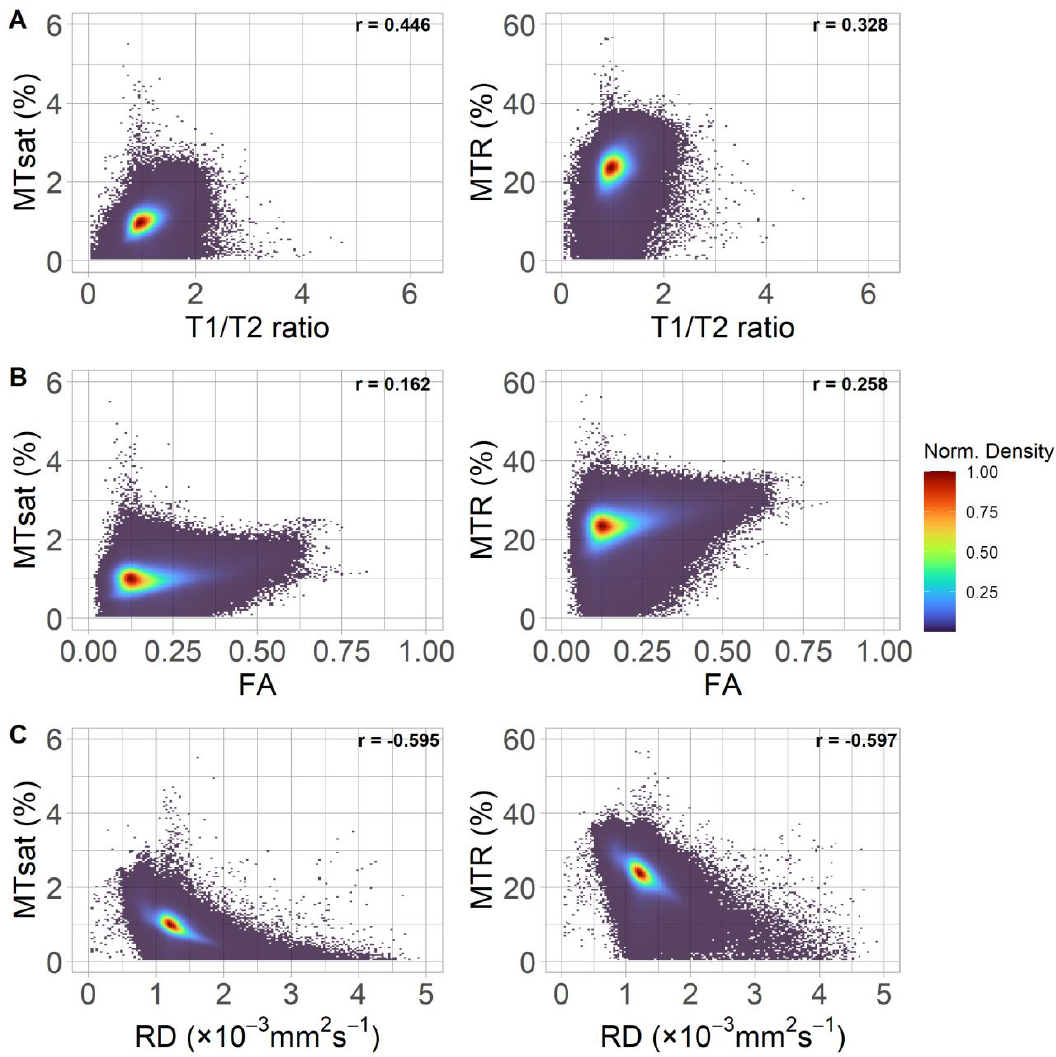
Two dimensional normalised density plots show the (binned voxelwise) relationship between the magnetisation transfer imaging metrics and: (A) T1w/T2w ratio, (B) FA, and (C) RD in white matter. Correlation coefficients presented are the repeated measures correlation calculated using rmcorr, with study participant as the repeated measure.

**Figure 5:**
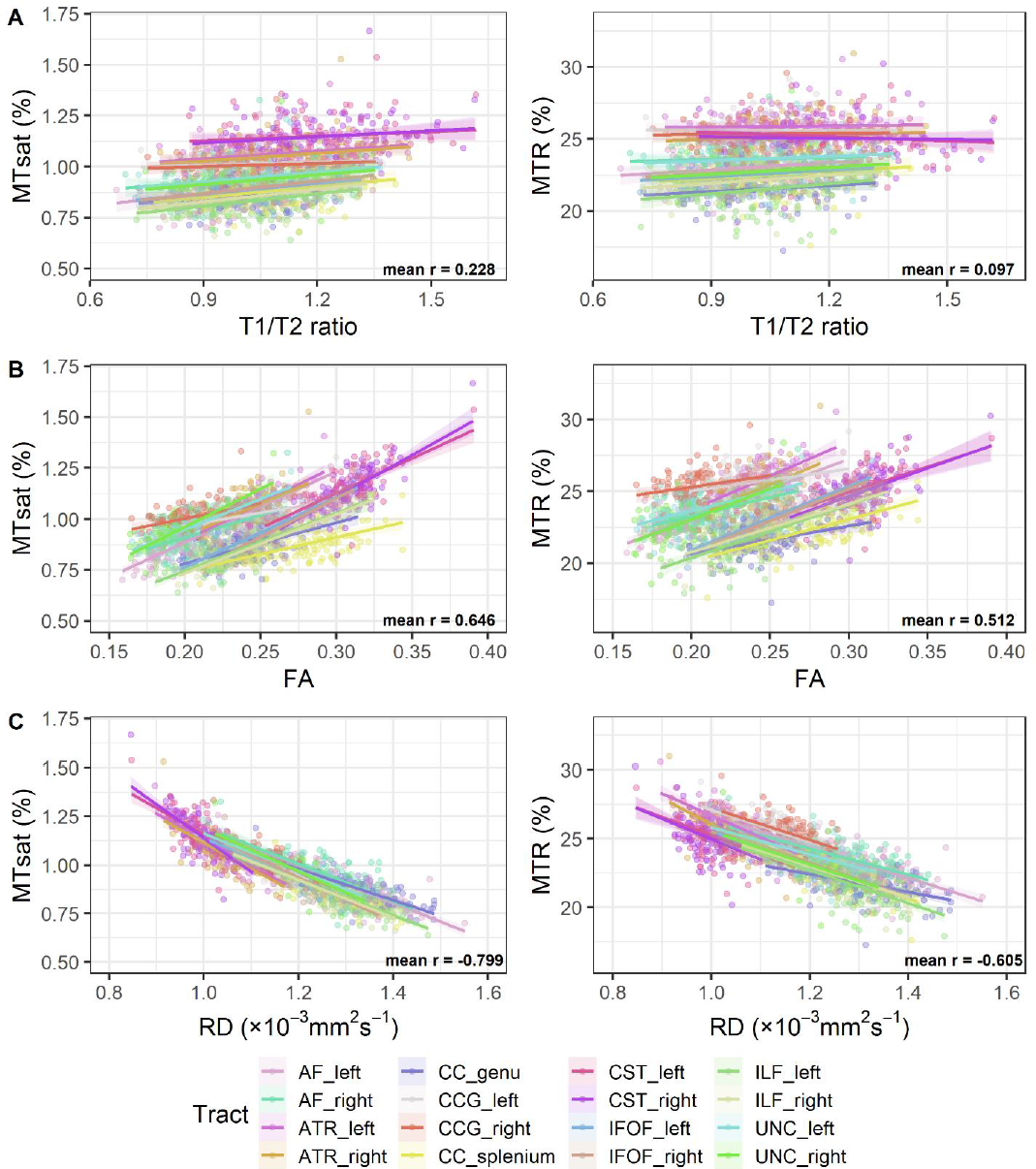
White matter tract-wise correlations between the two measures derived from magnetisation transfer imaging and: (A) T1w/T2w ratio (B) FA, and (C) RD. The average Pearson’s correlation coefficient for the relationships between the tracts was calculated by first transforming the Pearson’s r values to Fisher’s Z, taking the average, and then back-transforming the value to Pearson’s correlation coefficient.

## 4 Discussion

In this work, we used MTI to characterise myelination in the neonatal brain in association with age at scan and preterm birth. For the first time, MTsat was applied in a neonatal population. Across the WM, there were positive correlations between GA at scan and the MTI metrics. Preterm birth was associated with increased MTR in central WM regions and decreased MTsat in the genu and splenium of the corpus callosum. T1w/T2w ratio had moderate positive correlations with MTR and MTsat at voxel level, but weaker within major WM tracts, whilst the opposite was observed for FA. RD had the strongest negative correlations with both MTsat and MTR across WM voxels and major tracts. This study offers a new approach for myelin-sensitive imaging in early life, shows that MTI captures key features of EoP, and contributes to the understanding of how commonly used WM integrity measures relate to those more specific to myelin.

Both MTsat and MTR values in early life are remarkably lower than those reported in adult populations (York et al., 2022b). Average MTsat and MTR values in the WM of healthy adults are around 3.7% and 54.5%, respectively (York et al., 2022b), whilst in this neonatal sample, the range of average MTsat and MTR values in the major WM tracts are 0.8–1.18% and 21.5– 26.0%, respectively. These values are even lower than those observed in the WM lesions of multiple sclerosis patients (York et al., 2022b). This is likely to reflect lower myelin density in the neonatal compared to the adult brain. However, it also raises the question to what extent MTsat and MTR values in the neonatal brain are influenced by myelin density as compared with other biological processes. It is important to note that MTR is highly protocol dependent, making comparison of values across different studies difficult; the current study applied a MT protocol similar to the one used previously in the multiple sclerosis study (York et al., 2022b). Nevertheless, MTR values in a similar range (20–30%) have been reported previously in preterm neonates at TEA (Nossin-Manor et al., 2013).

Both MTsat and MTR are higher in the central (i.e. deep grey matter structures, brain stem and central white matter regions such as the posterior limb of the internal capsule [PLIC] - part of the corticospinal tract) compared to frontal/occipital regions, indicating higher myelin content in the centre of the brain. By quantifying the mean values across major WM tracts, we found that corticospinal tract and the anterior thalamic radiation had the highest level of myelination whilst the inferior longitudinal fasciculus and the genu/splenium of the corpus callosum were among the least myelinated tracts at TEA. Similar varying levels of myelination across WM regions in early infancy have been observed in other studies using different MRI techniques to measure myelination such as the T1w/T2w ratio (Filimonova et al., 2023; Grotheer et al., 2023; Soun et al., 2017), T1 (or its inverse R1) or T2 (or its inverse R2) mapping, (Grotheer et al., 2022; Kulikova et al., 2015; Leppert et al., 2009), or multi-component relaxometry to quantify myelin water fraction (Deoni et al., 2011; Melbourne et al., 2016, 2013). These studies have also reported features consistent with higher levels of myelination in the centre of the brain such as in the PLIC and lower levels in frontal and occipital regions, including the genu and splenium of the corpus callosum.

We observed strong positive correlations between GA at scan and MTR/MTsat across the WM, indicating increased myelination content with increasing age. These results are in line with previous studies showing higher MTR values in WM fibres with increasing age at scan during the neonatal period in preterm infants (Nossin-Manor et al., 2015, 2013). Furthermore, studies using other MRI methods to quantify myelin density in early life have found positive correlations with age at scan, both from preterm birth up to TEA as well as postnatally, with the fastest increase in myelin content happening between birth and first year of life. This includes methods such as calculation of T1w/T2w ratio (Filimonova et al., 2023; Grotheer et al., 2023; Lee et al., 2015; Thompson et al., 2022), T1 and T2 mapping (Counsell et al., 2003; Deoni et al., 2012; Grotheer et al., 2022; Leppert et al., 2009; Melbourne et al., 2016; Schneider et al., 2016), as well as myelin water fraction imaging (Dean et al., 2014; Deoni et al., 2012, 2011; Melbourne et al., 2016).Taken together, these data suggest that MTI is capturing the progression in myelin density that takes place during the developmental window period of data acquisition used in this study.

Decreased myelin content has been demonstrated in the preterm brain at TEA (Grotheer et al., 2023; Hagmann et al., 2009; Pannek et al., 2013) and this appears to persist throughout childhood (Thompson et al., 2022; Vandewouw et al., 2019), though regional effects vary between studies. Here, we found lower MTsat values in the preterm brain, with strongest effects in the genu and splenium of the corpus callosum and the inferior longitudinal fasciculus, reflecting lower myelination in these regions compared to term-born controls. Interestingly, these tracts had the lowest values of MTsat, suggesting that areas with lower myelination in the neonatal brain may be more affected by early exposure to extrauterine life. Previous studies have shown that regions with lower myelin content at birth, such as the genu and splenium of the corpus callosum, have the fastest increase in myelin density postnatally in the first year of life compared to regions with higher levels of myelin at birth, such as the corticospinal tract (Deoni et al., 2011; Filimonova et al., 2023; Grotheer et al., 2022). In contrast, studies of preterm infants from preterm birth up to TEA report that myelin density increases the fastest in the PLIC, whilst myelin imaging parameters change very little in the genu and splenium of the corpus callosum during this period (Melbourne et al., 2013; Nossin-Manor et al., 2013; Schneider et al., 2016); though some studies suggest linear increase in myelin MRI parameters in all white matter regions over this developmental time period (Counsell et al., 2003). Collectively, this suggests that preterm birth may have an effect on “late-myelinating” WM regions and that with MTsat this is already evident at TEA. Indeed, a recent study suggested that the rate of myelin development is more rapid *in utero* and slows down *ex utero*, leading to lower myelination in the preterm brain (Grotheer et al., 2023). Future studies are needed to ascertain if the lower MTsat values in the preterm WM persist into later developmental periods.

Counter-intuitively, we found higher MTR in preterm than term WM, particularly in tracts that had high MTR/MTsat values. However, it is important to remember that MTR is susceptible to T1 relaxation effects (Helms et al., 2008b). Importantly, none of the WM regions that showed higher MTR in the preterm WM had a correspondingly higher values of MTsat. Indeed, increase in cellular/axonal density and myelin-related macromolecules, paralleled with decreasing water content in the developing brain can result in increased MTR as well as R1 (Grotheer et al., 2022; Nossin-Manor et al., 2015; Yeatman et al., 2014). Whilst MTR has been shown to reflect myelin content in histological analysis in the adult brain (Mancini et al., 2020; Schmierer et al., 2004), to our knowledge, MTR has not been validated in terms of its correlation with histological myelin measurements in the neonatal brain. These results emphasise previous observations that caution is required when interpreting MTR data because of the sensitivity of R1 to a number of biological processes, such as cellular density, iron concentration, calcium content and axonal count and size which may have stronger contributions in the infant brain (Grotheer et al., 2022; Harkins et al., 2016). Thus, the effects of preterm birth observed for MTR may be driven by other factors besides myelin density.

The final aim of this study was to investigate the relationship between myelin-sensitive MTI metrics and commonly used neuroimaging markers of WM dysmaturation/integrity – T1w/T2w ratio, FA and RD. Previously, weak positive correlations have been reported between T1w/T2w ratio and MTsat (r=0.28 (Saccenti et al., 2020)) and strong correlations between T1w/T2w ratio and MTR (r=0.63 (Pareto et al., 2020)) in normal appearing WM in patients with multiple sclerosis. However, these relationships have not been studied in the neonatal brain. We observed moderate positive correlations between T1w/T2w ratio and MTR as well as MTsat in the WM at the voxel-level and weak positive correlations at tract-level. When investigating the correlations with FA, there was the opposite phenomenon as the one observed with T1w/T2w ratio: the average tract correlations within WM tracts are stronger than the voxel-wise correlations. Positive correlations between MTR and FA in WM regions at TEA have been shown previously (Nossin-Manor et al., 2015, 2013). Our results could potentially reflect different patterns of myelination across the brain. The general pattern of myelination relies on a caudo-rostral gradient, a progression from the brain centre to the periphery, in sensory and motor pathways before associative pathways (Dubois et al., 2021). This is reflected by our findings of correlations with differing magnitude between MTI metrics, and T1w/T2w ratio and FA across the WM tracts. FA is well-known to be affected by multiple factors based on the water content and geometry of the tracts (Figley et al., 2022), especially in crossing fibres areas, which represent around 90% of the brain (Jeurissen et al., 2013). This effect is less pronounced within the major WM tracts, where the geometry is simpler and the fibres are better/tightly aligned (Figley et al., 2022). This could explain why, on average, the correlation of FA with the MTI metrics is stronger in specific tracts compared to the whole WM voxel-wise approach. This finding may suggest that within major WM tracts, myelin could have a significant contribution to restricted water diffusion and that the development of axonal structure and myelin are closely coupled. On the other hand, the relatively high positive correlations of the T1w/T2w ratio with the MTI metrics at the whole WM level are highly reduced when looking at the tract level. This suggests that anatomical specificity is important in understanding the contribution of myelin to commonly used measures of WM integrity. However, the differences between tract- and voxel-level correlations between the metrics need to be investigated in future studies as it could be that some filtering should be applied to reduce noise in voxel-wise correlations.

Compared with FA and T1w/T2w ratio, RD had stronger negative correlations with MTsat and MTR. Negative correlation between RD and MTR has been shown in neonates previously (Nossin-Manor et al., 2015, 2013). The correlation magnitude was similar for MTsat and MTR across the WM voxels, whilst, similar to FA, in major WM tracts, RD had even stronger negative correlations with MTsat. Collectively, this finding illustrates strong correlations between RD and more myelin-sensitive imaging metrics, supporting previous literature that suggests RD sensitivity to myelin pathologies (Lazari and Lipp, 2021; Mancini et al., 2020; Song et al., 2002).

This study is the first to report MTsat values in a sample of neonates comprised of term-born controls and preterm infants without major parenchymal lesions i.e. a sample that is representative of the majority of survivors of neonatal intensive care. A multimodal acquisition protocol enabled co-registration of the MT images to diffusion space for delineation of major white matter tracts. This study has some limitations. We used a large voxel size of 2 mm^3^ for the MTI, but this was required to optimise acquisition parameters to achieve shorter acquisition times. Neonatal MT images are challenging to align to other modalities due to the low contrast between tissue characteristics of the neonatal brain (Dubois et al., 2021); to overcome this limitation, the T1w acquired during the MTI acquisition was co-registered to the T1w structural image, and this transformation was used as a bridge to move the maps from one space to another. Although calculation of MTsat inherently corrects for T1 relaxation and *B*_1_^+^ inhomogeneity effects (Helms et al., 2008b), correction for residual *B*_1_^+^ effects may still be needed (Rowley et al., 2021). Therefore, future studies which include *B*_1_^+^ map acquisitions are needed to ascertain whether *B*_1_^+^ correction of MTsat maps modifies interpretation of the main findings. It was beyond the scope of this study to investigate all available MT-based metrics; in future studies it would be useful to assess whether these results have histopathological correlates, and whether they are comparable to other quantitative MT-based indices such as the MPF (Kisel et al., 2022; Yarnykh, 2020).

Myelin imaging in infancy may provide novel biomarkers for neurodevelopmental outcomes later in childhood. For example, higher myelin density in infancy and early childhood, measured using MTI-derived MPF (Corrigan et al., 2022; Zhao et al., 2022), T1w/T2w ratio (Darki et al., 2021), or myelin water fraction (Dai et al., 2019; Deoni et al., 2016; O’Muircheartaigh et al., 2014), correlate with improved cognitive outcomes such as performance in executive function tasks and language skills. Furthermore, T1w/T2w ratio and T2 relaxometry values in 1-9 month old infants have been associated with familial risk of autism spectrum disorder (Darki et al., 2021) and cerebral palsy (Chen et al., 2018), respectively. To our knowledge, only one study has investigated the predictive value of myelin MRI measures at TEA for later outcomes, finding no significant relationships with T1w/T2w ratio at TEA, though significant associations were demonstrated at 7 and 13 years of life with a range of cognitive scores (Thompson et al., 2022). This suggests that there is uncertainty to what extent variation in neonatal myelin imaging metrics, including MTsat, associates with neurodevelopmental outcomes in childhood. The participants in this study are part of a longitudinal cohort, which provides opportunity in the future to assess relationships between neonatal MTI and functional outcomes in childhood.

## 5 Conclusions

This study provides a new characterisation of the neonatal brain using MTI, and demonstrates the utility of the technique for studying disorders of myelination in early life. Both MTsat and MTR increase with GA at scan. In term compared with preterm infants, MTsat is higher while MTR is lower. This could suggest that MTsat may be a more reliable biomarker of myelin in the neonatal brain, and cautions the use of MTR to measure myelin density due to the confounding effects of R1/T1. In addition, by correlating MTI metrics with common WM integrity biomarkers, FA, RD and the T1w/T2w ratio, we observed interesting opposing trends at voxel- and tract-level, which emphasises the necessity to incorporate anatomical information when interpreting the contribution of myelin to non-specific imaging metrics in early life studies. Future studies will investigate the utility of preterm birth-associated differences in neonatal MTsat in terms of their relevance to neurodevelopmental and cognitive outcomes.

## 6 CRediT authorship statement

**Manuel Blesa Cábez**: Conceptualization, Methodology, Software, Formal analysis, Data Curation, Writing - Original Draft, Visualization; **Kadi Vaher**: Conceptualization, Methodology, Formal analysis, Investigation, Data Curation, Writing - Original Draft, Visualization; **Elizabeth N. York**: Formal analysis, Software, Writing - Review & Editing; **Paola Galdi**: Formal analysis, Writing - Review & Editing; **Gemma Sullivan**: Investigation, Data Curation; **David Q. Stoye**: Investigation, Data Curation, Writing - Review & Editing; **Jill Hall**: Data Curation, Project administration; **Amy E. Corrigan**: Investigation, Data Curation; **Alan J. Quigley**: Investigation; **Adam D. Waldman**: Writing - Review & Editing; **Mark E. Bastin**: Methodology, Software, Resources, Writing - Review & Editing; **Michael J. Thrippleton**: Methodology, Software, Resources, Writing - Review & Editing; **James P. Boardman**: Conceptualization, Methodology, Writing - Original Draft, Supervision, Funding acquisition.

## Data Availability

Requests for anonymised data will be considered under the study's Data Access and Collaboration policy and governance process (https://www.tebc.ed.ac.uk/2019/12/data-access-and-collaboration/).

## Abbreviations

AF: arcuate fasciculus
ATR: anterior thalamic radiation
CC genu: corpus callosum genu/forceps minor
CC splenium: corpus callosum splenium/forceps major
CCG: cingulum cingulate gyrus
CST: corticospinal tract
dHCP: developing human connectome project
dMRI: diffusion magnetic resonance imaging
FA: fractional anisotropy
FDR: false discovery rate
FWER: family-wise error correction
GA: gestational age
GM: grey matter
IFOF: inferior fronto-occipital fasciculus
ILF: inferior longitudinal fasciculus
MPF: macromolecular proton fraction
MRI: magnetic resonance imaging
MTI: magnetisation transfer imaging
MTR: magnetisation transfer ratio
MTsat: magnetisation transfer saturation
R1app: approximation of R1
RD: radial diffusivity
ROI: region of interest
SNR: signal-to-noise ratio
TE: echo time
TEA: term-equivalent age
TFCE: threshold-free cluster enhancement
TR: repetition time
UNC: uncinate fasciculus
WM: white matter

## 8 Acknowledgments

This research was funded in whole, or in part, by the Wellcome. For the purpose of open access, the author has applied a CC BY public copyright licence to any Author Accepted Manuscript version arising from this submission. We are grateful to the families who consented to take part in the study. Neonatal participants were scanned in the University of Edinburgh Imaging Research MRI Facility at the Royal Infirmary of Edinburgh which was established with funding from The Wellcome Trust, Dunhill Medical Trust, Edinburgh and Lothians Research Foundation, Theirworld, The Muir Maxwell Trust and many other sources. We are thankful to all the University’s imaging research staff for providing the infant scanning.

