## Supplementary Figure for "Characterisation of the neonatal brain using myelin-sensitive magnetisation transfer imaging"

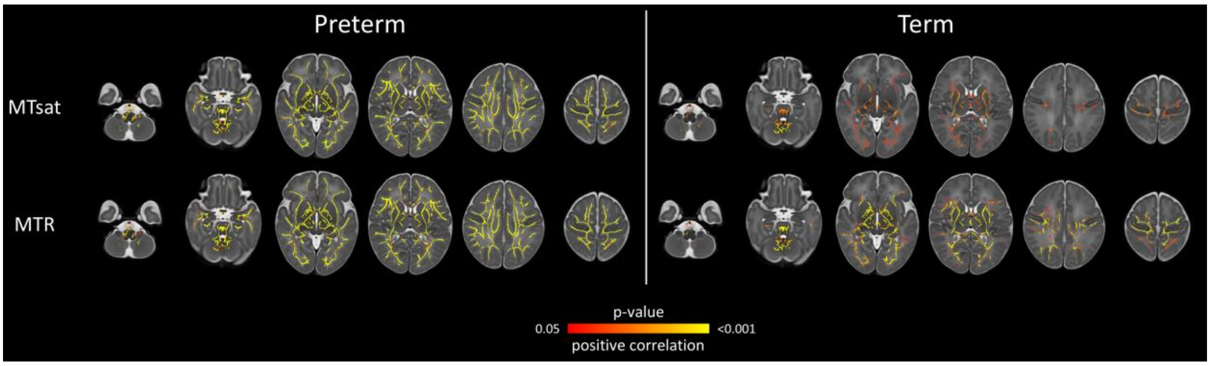


*Supplementary Figure 1: Voxel-wise analysis showing effects of GA at scan on magnetisation transfer imaging metrics separately in preterm (left) and term (right) groups (see main paper Figure 2 for the skeleton mask). Models were adjusted for GA at birth. Voxels that have positive correlation with GA at scan are indicated in red-yellow, overlaid on the dHCP T2w 40-week template. Results are reported after 5000 permutations, p-values corrected using TFCE and FWE with a significance level of p<0.05. For visualisation: anatomic left is on the right side of the image. GA = gestational age, FA = fractional anisotropy, RD = radial diffusivity, FWE = family-wise error correction, TFCE = threshold-free cluster enhancement.*

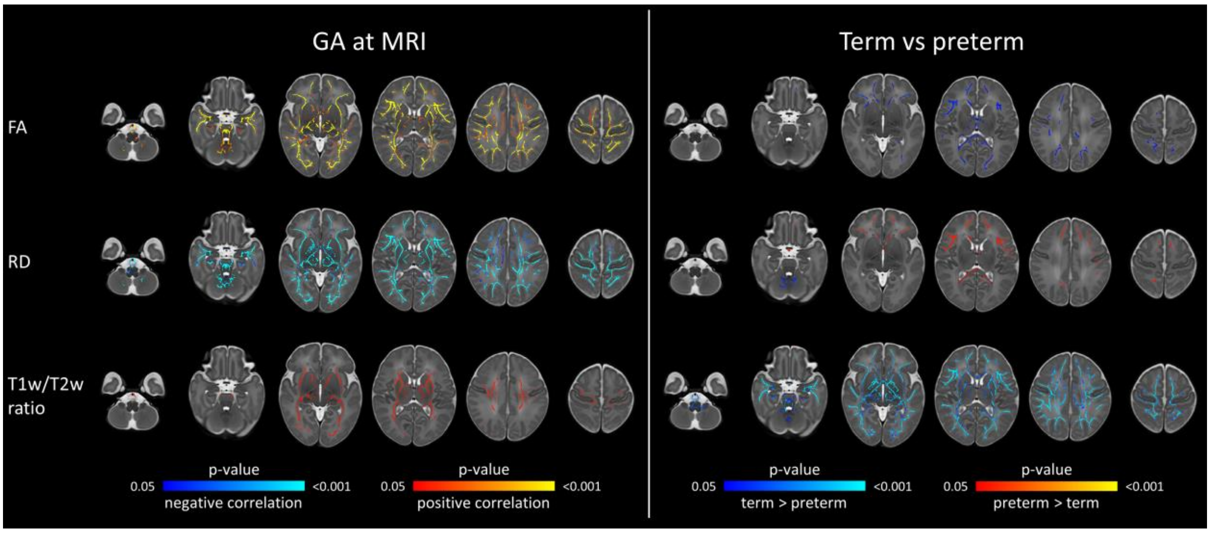


*Supplementary Figure 2: Voxel-wise analysis showing effects of GA at scan and preterm birth on diffusion tensor imaging metrics and the T1w/T2w ratio in the white matter skeleton (see main paper Figure 2 for the skeleton mask). Models were mutually adjusted for GA at scan and preterm status. In left panel, voxels that have positive correlation with GA at scan are indicated in red-yellow, and those with negative correlation are indicated in blue-light blue. In right panel, voxels that have higher values in preterm compared with term group are indicated in red-yellow; voxels that have higher values in term compared with preterm group are indicated in blue-light blue. Overlaid on the dHCP T2w 40-week template. Results are reported after 5000 permutations, p-values corrected using TFCE and FWE with a significance level of p<0.05. For visualisation: anatomic left is on the right side of the image. GA = gestational age, FA = fractional anisotropy, RD = radial diffusivity, FWE = family-wise error correction, TFCE = threshold-free cluster enhancement.*
